# Statistical Properties of Stepped Wedge Cluster-Randomized Trials in Infectious Disease Outbreaks

**DOI:** 10.1101/2020.05.01.20087429

**Authors:** Lee Kennedy-Shaffer, Marc Lipsitch

## Abstract

Randomized controlled trials are crucial for the evaluation of interventions such as vaccinations, but the design and analysis of these studies during infectious disease outbreaks is complicated by statistical, ethical, and logistical factors. Attempts to resolve these complexities have led to the proposal of a variety of trial designs, including individual randomization and several types of cluster randomization designs: parallel-arm, ring vaccination, and stepped wedge designs. Because of the strong time trends present in infectious disease incidence, however, methods generally used to analyze stepped wedge trials may not perform well in these settings. Using simulated outbreaks, we evaluate various designs and analysis methods, including recently proposed methods for analyzing stepped wedge trials, to determine the statistical properties of these methods. While new methods for analyzing stepped wedge trials can provide some improvement over previous methods, we find that they still lag behind parallel-arm cluster-randomized trials and individually-randomized trials in achieving adequate power to detect intervention effects. We also find that these methods are highly sensitive to the weighting of effect estimates across time periods. Despite the value of new methods, stepped wedge trials still have statistical disadvantages compared to other trial designs in epidemic settings.

---

Randomized controlled trials are crucial to evaluating interventions including vaccines and other preventative measures during infectious disease outbreaks. Epidemic settings and vaccine studies however pose statistical logistical and ethical challenges that make randomized trials more difficult to design conduct and analyze (1). Statistically, trials must account for the interference between individuals and define explicitly whether they identify the direct or indirect effects of the vaccine, as well as handle a high degree of spatiotemporal variation, uncertain incidence rates, and the potential for mild or asymptomatic infections (2–4). Logistically, trials must be able to be implemented in the context of ongoing epidemiological work in outbreaks and account for the timeline of production of the vaccine and the speed at which it can be rolled out to affected communities (3,5,6). Ethically, vaccine trials face complex considerations of the overall value of the trial as well as the risks and benefits to participants and individuals in communities with participants (7,8).

Cluster-randomized trials (CRTs) have recently become more common in infectious disease settings. These designs are well-suited to capture indirect effects (e.g., the effects of herd immunity) and, in some situations, may be logistically easier to implement or more acceptable to participating communities (3,7,9). More complex CRT designs have also been proposed for vaccine trials in outbreak settings. These include the ring vaccination design that was used in the Ebola outbreak in Guinea in 2015 (10), as well as the stepped wedge cluster-randomized trial (SWT) design, which has been proposed in various outbreak settings, including the Ebola outbreak in Sierra Leone (11). SWTs may be more acceptable to communities enrolling in trials because there is no placebo group, and they may align well with a phased rollout necessitated by implementation challenges (12–13).

There are, however, tradeoffs to these designs. They are not designed to identify direct effects of intervention (7). In addition, CRTs and SWTs generally have lower power to detect treatment effects and thus require a larger sample size than individually-randomized trials (IRTs) (9,14,15). They also may exhibit biases due to imbalance between clusters, especially with the high incidence heterogeneity of outbreaks, and they have less flexibility to adapt the design or increase sample size (3,9,11,16). To better understand the statistical properties of these designs, we can evaluate their performance on simulated outbreaks (17). Bellan et al. found that SWTs had much lower power than IRTs in simulations of the waning Ebola outbreak (14). Hitchings et al. used simulated outbreaks to examine the tradeoff between capturing indirect vaccine effects and reduced power between parallel-arm CRTs and IRTs (15).

SWTs in particular are highly susceptible to misspecification and can produce biased results when time trends and time-intervention interactions are not modeled correctly (18,19). Type I Error of hypothesis tests can be preserved by using permutation-based inference, but this generally results in reduced power (20–22). New methods for analyzing SWTs have recently been proposed which preserve Type I Error but may have more precision and higher power than permutation tests based on mixed effects models when applied to data-generating settings for which mixed effects models are misspecified. These can be purely “vertical” methods that avoid the need to model time trends, like the non-parametric within-period method (23), the design-based approach (24), and the synthetic control-based approach (22), or “horizontal” methods that compare within-cluster differences between two time points across clusters (22).

The properties of these analysis methods have been studied in various theoretical and simulation-based contexts, but not specifically for infectious disease outbreaks, and not in a context that compares them to IRT and CRT designs.

Understanding the statistical properties of various trial designs for infectious disease outbreaks is a key part of planning for vaccine studies. Vaccine studies in outbreak settings, like COVID-19, should take into consideration these properties, along with feasibility and ethical considerations, in the design phase. To be ethical, a randomized trial should have a clear analysis plan that will result in a statistically valid estimate of the effect and is adequately powered to detect a meaningful effect size in a reasonable amount of time. By considering the properties of various SWT analysis methods and comparing these to IRT and CRT methods, this article contributes to the appropriate design of future trials conducted during epidemics.

## METHODS

### Outbreak and trial simulation

We simulate outbreaks using a model developed by Hitchings et al. (15). This simulates a main population, in which an epidemic progresses, and the study population, which is comprised of many smaller communities. Infections are imported from the main population into the study population, where the outbreak spreads within the communities, but, for our simulation, not between communities.

The model and parameters are described in more detail by Hitchings et al. (15). We use the infectious period distribution (Gamma distributed with a mean of 5.0 days with a standard deviation of 4.7 days), community size (uniformly distributed from 80 to 120), within-community probability of a contact between two individuals (0.15), and percentage of a community enrolled in the trial (50%) used in those simulations. To reduce the number of communities with no cases, we increase the expected number of importations into a community to two over the course of the study. We assume the incubation period and latent period are the same for each individual. The length of this period is independently generated for each individual from a Gamma distribution with shape parameter 5.807 and scale parameter 0.948, for a mean incubation period of 5.51 days, as has been estimated for COVID-19 (25). We enroll 40 communities into the trial 56 days after the start of the epidemic in the main population and conduct follow-up for 308 days. Finally, we consider four values of R_0_ for the outbreak: R_0_ = 1.34, 1.93, 2.47, and 2.97, by varying the transmission rate constant parameter in the model.

On top of this outbreak, we simulate three types of randomized trials: an individually-randomized trial (IRT), a CRT, and a SWT. The IRT and CRT are conducted as described by Hitchings et al. (15). In all designs, on day 56, half of the individuals in each study cluster who have not yet been infected are enrolled into the trial. In the IRT, half of these individuals in each cluster are assigned to vaccination and the other half to control; the vaccination occurs immediately upon enrollment. In the CRT, half of the clusters are assigned to vaccination and the other half to control; all enrolled individuals in a cluster receive the treatment for that cluster immediately upon enrollment. In the SWT, all clusters begin in the control arm. In design SWT-A, four clusters cross over to the vaccination arm every 28 days, beginning on day 84. In design SWT-B, one cluster crosses over to the vaccination arm every 7 days, beginning on day 84. We consider three values of the direct vaccine efficacy: VE=0, 0.60, and 0.80. In both of the non-zero cases, the vaccine is leaky, conferring a constant reduction in the probability of infection acquisition per contact across all vaccinated individuals.

### Analysis methods

The results from the IRT and CRT designs are analyzed as described by Hitchings et al., using the time to symptom onset for each enrolled individual (15). For the IRT, statistical analysis is conducted using a stratified Cox proportional hazards analysis, stratified by community. For the CRT, statistical analysis is conducted using a Cox proportional hazards model with a gamma-distributed shared frailty to account for clustering by community.

The results from the two SWT designs are analyzed using a variety of methods, all based on the number of individuals with symptom onset in each cluster in each period. This method has the advantage of being less sensitive to consistent determination of the date of symptom onset, but may have less power than methods using the time to event. The methods used are:

- MEM: Mixed effects model with a fixed effect of time and a normally-distributed random effect for cluster, with a logit link (26);
- CPI: Mixed effects model with a fixed effect of time and independent normally distributed random effects for cluster and cluster-period, with a logit link (27);
- Two vertical non-parametric within-period methods, both with a log link (23):
  - NPWP-1: Equally weighting period-specific NPWP estimates across periods;
  - NPWP-2: Weighting period-specific NPWP estimates by the total number of cases in that period;
- Four vertical synthetic control methods, all with a log link (22):
  - SC-1: Equally weighting clusters within each period and equally weighting period-specific SC estimates across periods;
  - SC-2: Equally weighting clusters within each period and weighting period-specific NPWP estimates by the total number of cases in that period;
  - SC-Wt-1: Weighting clusters within each period by the inverse mean square prediction error (MSPE) of the synthetic control fit and equally weighting period-specific SC estimates across periods;
  - SC-Wt-2: Weighting clusters within each period by the inverse MSPE of the synthetic control fit and weighting period-specific NPWP estimates by the total number of cases in that period.

We do not consider the horizontal crossover method since it is not well suited to capture indirect effects and is highly sensitive to time trends (22). To account for the delay in symptom onset (and thus the delayed effect of the intervention), we remove the first period on intervention for each cluster from all SWT analyses. For IRT and CRT analyses, we remove any infections that occur within the first six days (the average incubation time) of the trial.

For NPWP and SC, we take averages of the intervention cluster outcomes and control (or synthetic control) cluster outcomes within each period and then apply the contrast function, to reduce the effect of clusters with zero cases. For periods with zero cases among either the control clusters or intervention clusters, we add one-half case and one-half non-case to each arm. Periods with zero cases in both arms do not contribute to the effect estimate. For hypothesis testing, we use asymptotic inference for MEM and CPI and permutation inference for NPWP and SC (20–22). Permutation inference was not done for the MEM and CPI methods because of the high computational burden of these methods.

## RESULTS

### VE estimates and power by analysis method

We show main results for R_0_ = 2.47 and direct vaccine efficacy of VE = 0.6 for the IRT, CRT, and SWT-A (panel A) and SWT-B (panel B) designs. Figure 1 shows the median and first and third quartiles of the vaccine effectiveness estimates across 500 simulations. These results demonstrate that the IRT has the least variability among estimates, and is centered near the true direct vaccine efficacy. The CRT estimates a higher effectiveness, as it captures some indirect effects, but with higher variability. The SWT results are very dependent on the analysis method chosen, but all have higher variability than the CRT results and have a lower median estimate. Among SWT results, a higher effect is estimated when weighting across periods by the total number of cases in a given period than weighting equally. A higher effect is generally also estimated for SWT-A than for SWT-B.

**Figure 1.**
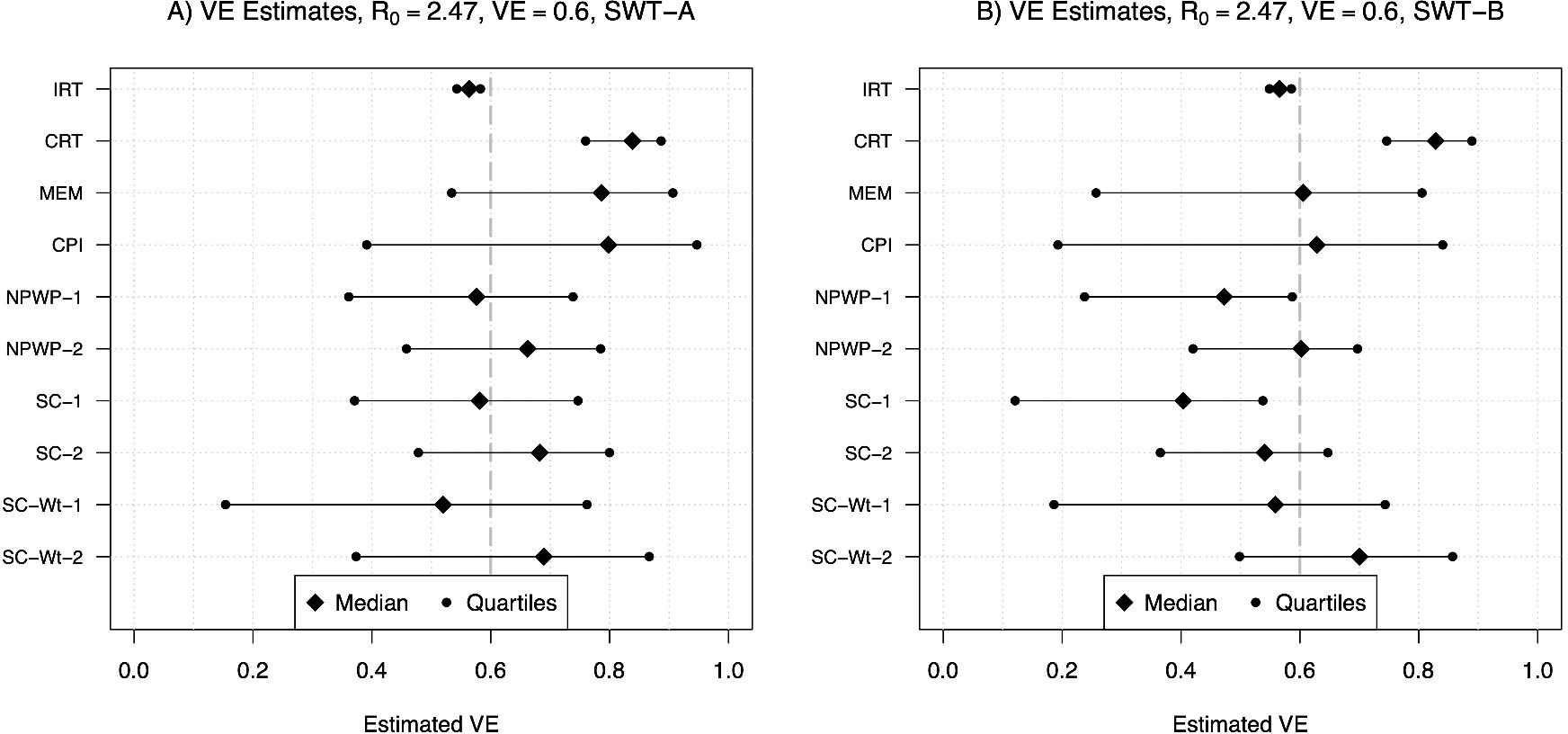
Median, first quartile, and third quartile of VE estimates for direct vaccine effect of 0.6 for individually-randomized trial analyzed with stratified Cox model (IRT), cluster-randomized trial analyzed with a Cox model with a gamma-distributed shared frailty (CRT), and stepped wedge trials with four clusters crossing over every 28 days (SWT-A, panel A) and with one cluster crossing over every 7 days (SWT-B, panel B) analyzed by mixed effects model (MEM), mixed effects model with cluster-period random effect (CPI), non-parametric within-period method equally weighted across periods (NPWP-1) and weighted across periods by total case count (NPWP-2), and synthetic control method equally weighted across clusters and periods (SC-1), equally weighted across clusters and weighted across periods by total case count (SC-2), weighted across clusters by inverse MSPE and equally weighted across periods (SC-Wt-1), and weighted across clusters by inverse MSPE and across periods by total case count (SC-Wt-2).

Figure 2 shows the power (panel A, VE = 0.6) and Type I Error (panel B, VE = 0) for asymptotic inference for the IRT, CRT, MEM, and CPI analyses and permutation inference for the NPWP and SC analyses. As seen in other settings, the asymptotic inference for MEM and CPI leads to greatly inflated Type I Error (14,20,22). The permutation inference for NPWP and SC has greatly reduced power compared to the IRT and CRT methods, with less than 50% power to detect a true direct vaccine effect of 0.6 compared to over 90% for the CRT and nearly 100% for the IRT. The NPWP-2 method achieves greater power in SWT-B than in SWT-A, but this is not the case for NPWP-1. SC-1 and SC-2 perform comparably to NPWP-1 and NPWP-2, respectively, but the SC-Wt methods have noticeably lower power.

**Figure 2.**
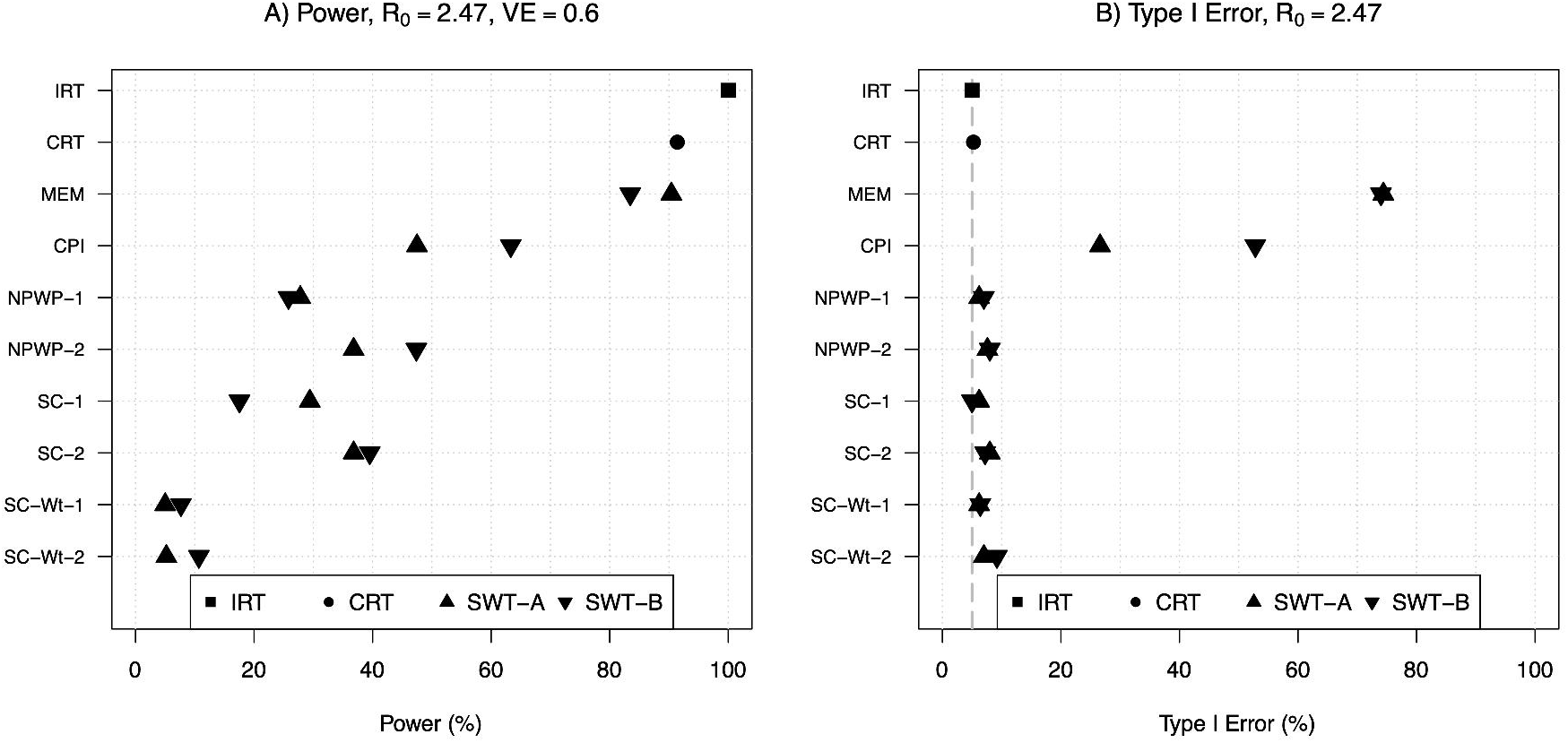
Empirical power for direct vaccine effect of 0.6 (panel A) and Type I Error (panel B) for individually-randomized trial analyzed with stratified Cox model (IRT), cluster-randomized trial analyzed with a Cox model with a gamma-distributed shared frailty (CRT), and stepped wedge trials with four clusters crossing over every 28 days (SWT-A, panel A) and with one cluster crossing over every 7 days (SWT-B, panel B) analyzed by mixed effects model (MEM), mixed effects model with cluster-period random effect (CPI), non-parametric within-period method equally weighted across periods (NPWP-1) and weighted across periods by total case count (NPWP-2), and synthetic control method equally weighted across clusters and periods (SC-1), equally weighted across clusters and weighted across periods by total case count (SC-2), weighted across clusters by inverse MSPE and equally weighted across periods (SC-Wt-1), and weighted across clusters by inverse MSPE and across periods by total case count (SC-Wt-2).

### Vaccine effectiveness estimates and power by R_0_

Figure 3 demonstrates the effect of R_0_ on the median VE estimate and observed power among these methods for the true direct VE = 0.6. Figure 4 shows the same results for true direct VE = 0.8. For both VE values, as R_0_ increases, both the estimated VE and the power of all of the SWT-A and SWT-B methods decrease. The same trend occurs for the CRT, although it maintains nearly 100% power when VE = 0.8 for all R_0_ values considered here. The IRT approach maintains its estimate and power throughout. The SWT methods decrease much more quickly than the CRT method, although there is no noticeable difference in this regard among the various SWT methods. For higher R_0_ values, the epidemic is passing so quickly through the communities that many communities have already experienced the epidemic before crossing over to the intervention, thus reducing the power of the SWT methods to detect effects. Throughout, the SC and NPWP methods perform similarly. While these figures only display NPWP-2 and SC-2, the same trends hold for NPWP-1 and SC-1. Similar trends, but with lower power throughout, hold for SC-Wt-1 and SC-Wt-2. Time-varying vaccine effects and weighting of vertical SWT methods

**Figure 3.**
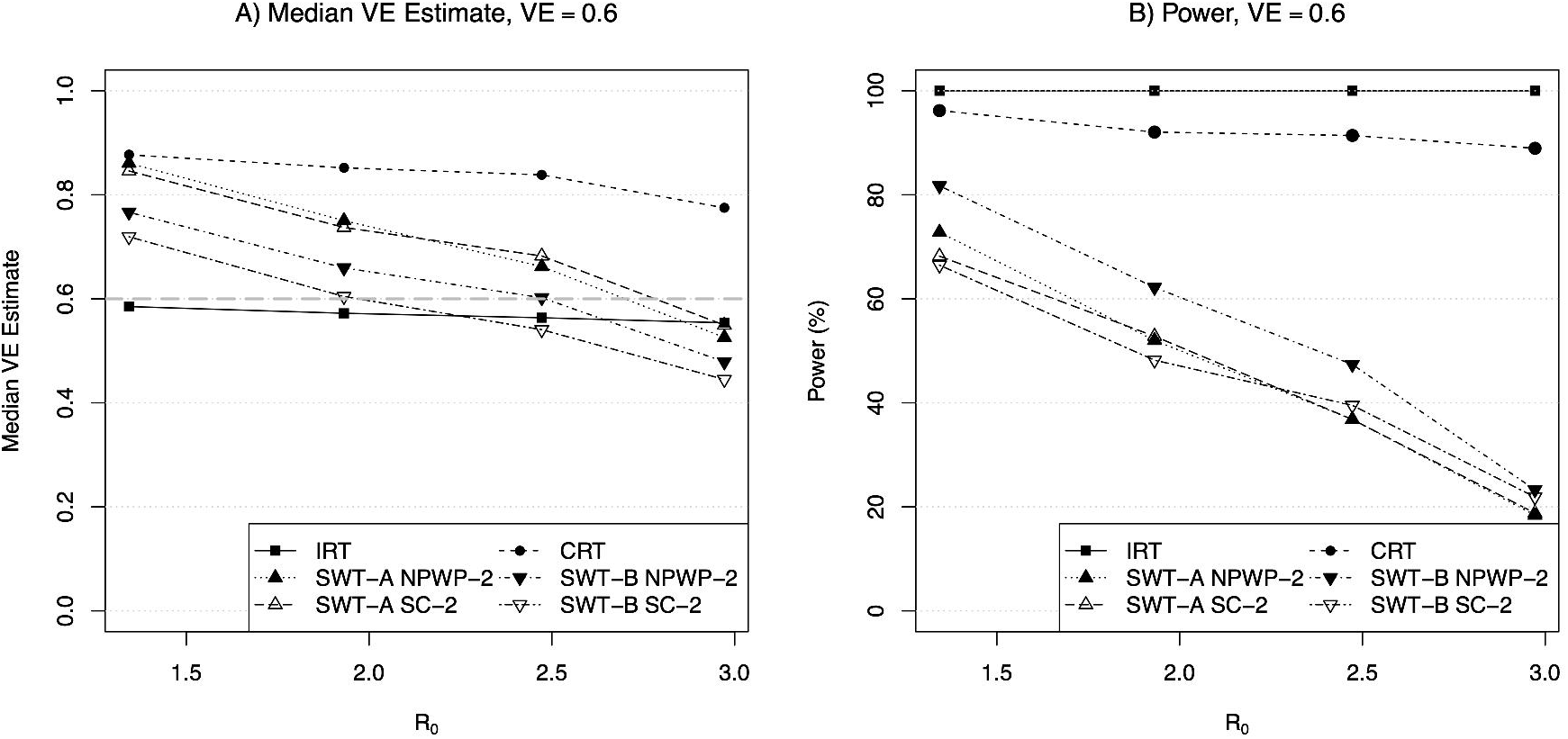
Median VE estimate (panel A) and empirical power (panel B) for direct vaccine effect of 0.6 vs. R_0_, for individually-randomized trial analyzed with stratified Cox model (IRT), cluster-randomized trial analyzed with a Cox model with a gamma-distributed shared frailty (CRT), and stepped wedge trials with four clusters crossing over every 28 days (SWT-A) and with one cluster crossing over every 7 days (SWT-B) analyzed by non-parametric within-period method weighted across periods by total case count (NPWP-2) and synthetic control method equally weighted across clusters and weighted across periods by total case count (SC-2).

**Figure 4.**
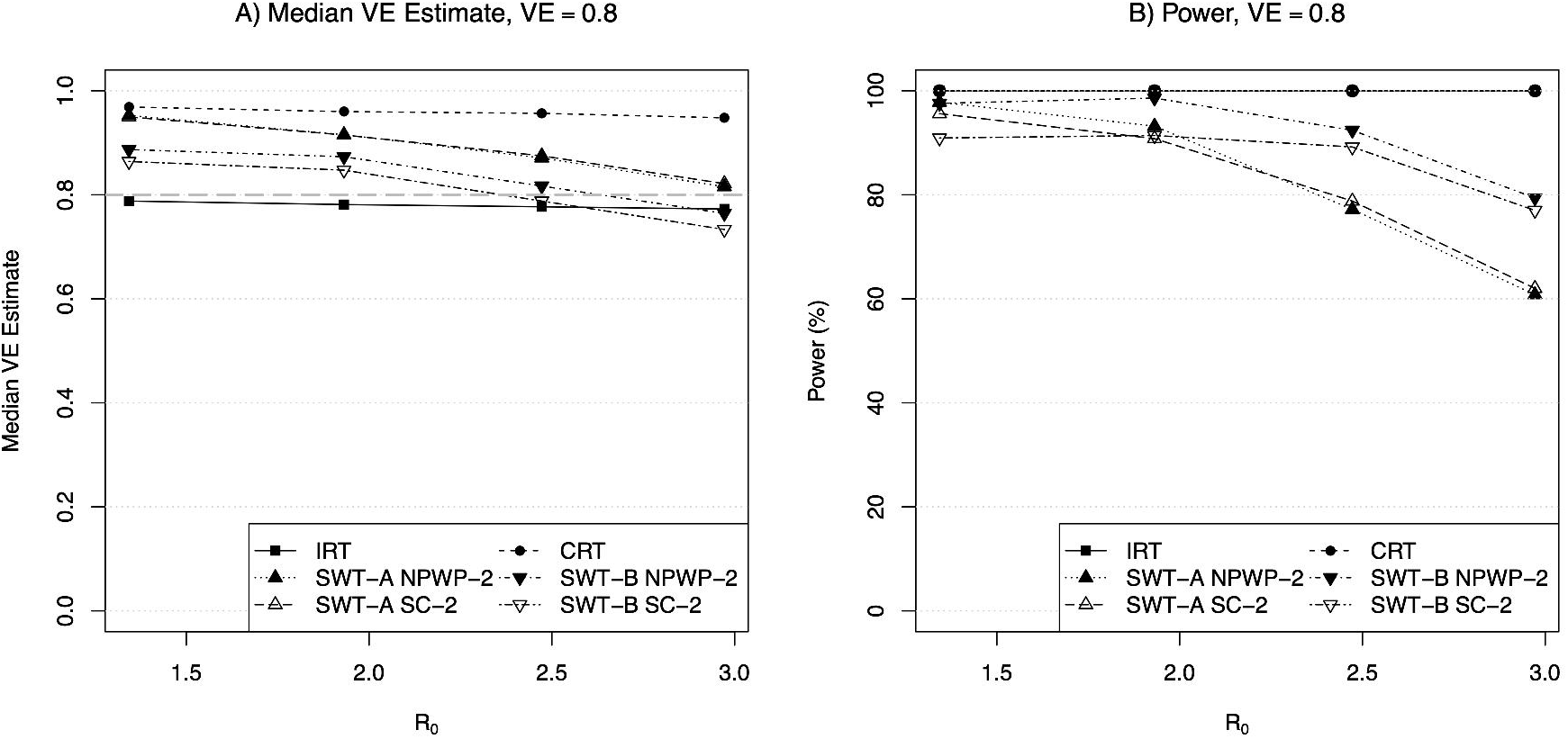
Median VE estimate (panel A) and empirical power (panel B) for direct vaccine effect of 0.8 vs. R_0_, for individually-randomized trial analyzed with stratified Cox model (IRT), cluster-randomized trial analyzed with a Cox model with a gamma-distributed shared frailty (CRT), and stepped wedge trials with four clusters crossing over every 28 days (SWT-A) and with one cluster crossing over every 7 days (SWT-B) analyzed by non-parametric within-period method weighted across periods by total case count (NPWP-2) and synthetic control method equally weighted across clusters and weighted across periods by total case count (SC-2).

The vertical methods for analysis of SWTs allow the investigator the ability to specify the weighting across periods considered in the study as well as, to some extent, weighting among clusters within each period. These choices can have a substantial effect on the overall estimated effect, as well as the power of the analysis. Figure 5 displays the estimated period-specific treatment effect (on the VE scale) for SWT-A (panel A) and SWT-B (panel B), analyzed by NPWP, SC, and SC-Wt. In both panels, for all three methods, there is a clear trend of maximum effect estimate early in the trial (although not at the very beginning for SWT-B) and a declining effect as the trial continues. For SWT-B, the negative effect estimates early in the trial are likely due to the very small number of clusters on intervention at that point, which can lead to a few simulations with a high number of early cases in those clusters having a big effect on the averages presented here. Later in the trial, clusters that have been on control throughout are more likely to have exhausted the susceptible population than clusters already intervened-upon, leading to lower incidence in the control clusters than the intervention clusters at that point. In other words, the flattening of the epidemic curve due to the intervention leads to an apparent decreased effectiveness of the intervention in later periods as the intervention clusters still have more remaining susceptible individuals than the control clusters.

**Figure 5.**
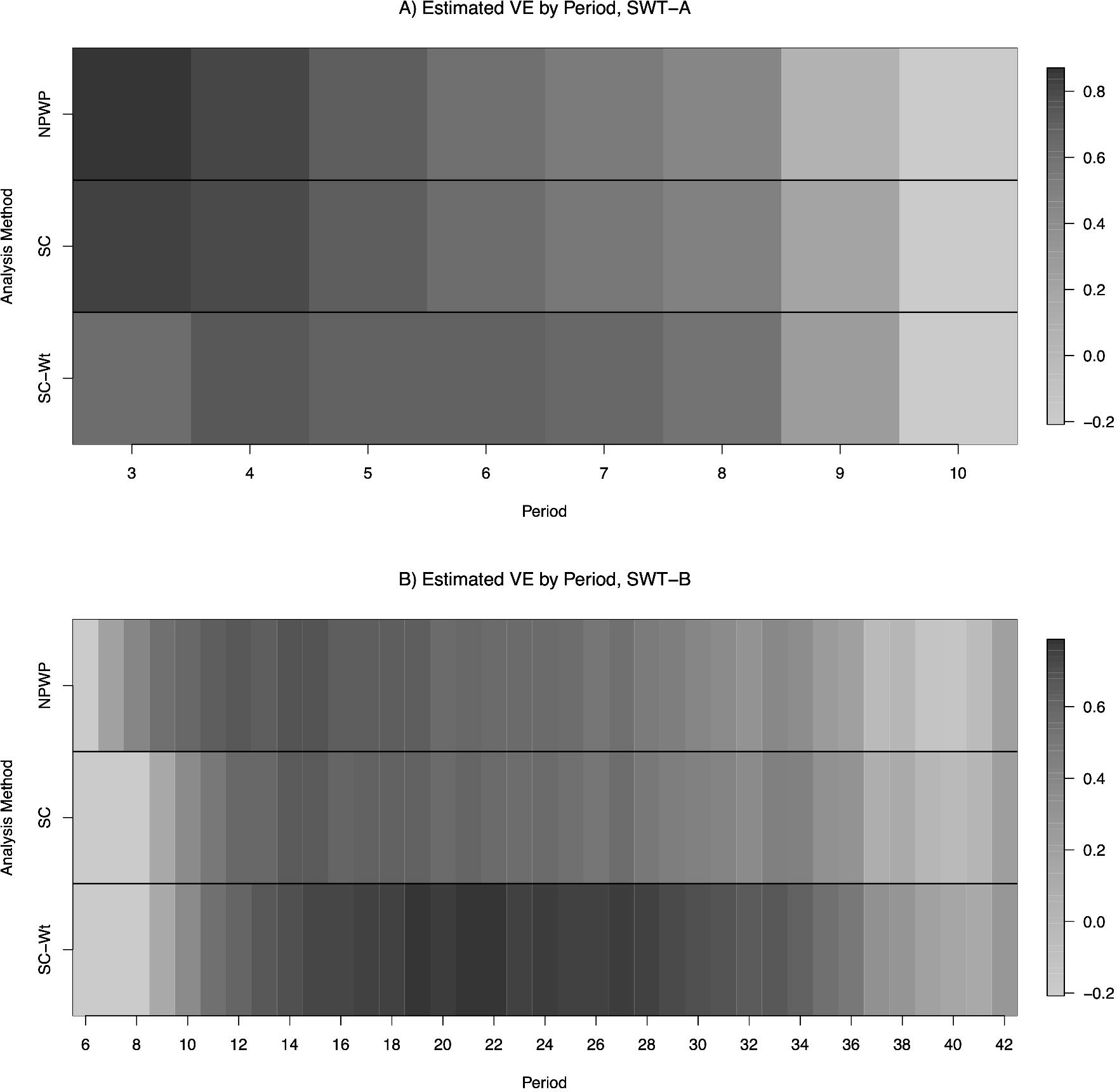
Average period-specific VE estimate by period for stepped wedge trials with four clusters crossing over every 28 days (SWT-A, panel A) and one cluster crossing over every 7 days (SWT-B, panel B) with VE = 0.6 and R_0_ = 2.47, analyzed by non-parametric within-period method (NPWP), synthetic control method weighted equally across clusters (SC) and synthetic control method weighted across clusters by inverse MSPE (SC-Wt).

While this freedom to target effect estimates to specific time scales could be useful for certain scientific questions, it also allows more researcher degrees of freedom in the analysis. Results of an SWT will thus be inherently limited by the duration and timing of the trial, decreasing the generalizability of these results. It may be difficult to select the optimal weighting method *a priori* as it likely depends on the temporal variation of incidence in the setting under study.

## DISCUSSION

The statistical performance of the SWT analysis methods considered here in simulated outbreaks highlights the drawbacks of the SWT design for the assessment of vaccines and other preventative measures during infectious disease outbreaks. Because of the high spatiotemporal variance of outbreaks, mixed effects model analyses of SWTs have greatly inflated Type I Error. This is especially true when the outbreak has a high R_0_, resulting in rapid spread within communities. And while permutation inference of purely vertical analysis methods (like the non-parametric within-period method and synthetic control method) can preserve Type I Error, they have greatly reduced power compared to analysis methods for other trial designs.

Additionally, the results of these vertical analysis methods are very dependent on the weighting scheme used to combine period-specific estimates and on the time period duration. In particular, the time trend in the number of intervention and control clusters removes the overall exchangeability of these groups and results in time-dependent cluster-level intervention effects. This weakens a key advantage of randomization: that it ensures comparability between the two groups (3). While the design may provide useful information on the relative effects of intervening at different points in the outbreak, it provides less clear evidence on the overall efficacy of the intervention, and the generalizability of the results may suffer. More research is needed to understand the trends in the estimated effect and power to detect an effect as the timing and duration of a CRT or SWT vary.

These simulated results focus narrowly on the statistical properties of the design. The ethical concerns, including the effect on trial participants and the speed with which a conclusion is reached, are also crucial considerations (3,7,28,29). In addition, the logistics of implementing the intervention may limit the choices available to trial designers. However, these issues may be better solved with risk prioritization in IRT or parallel-arm CRT designs rather than SWTs (14,28,29). All of these factors should be considered and appropriately weighed when designing a trial.

Further research is needed to further elucidate the relative advantages of various designs and analysis methods when a trial starts at different points relative to the outbreak curve. In addition, future work may consider the ability of methods to determine the relative benefits of beginning and ending interventions at different points. The stepped wedge design may be useful for that purpose, as it can provide information on the time trends in intervention effectiveness, but other designs may be valuable for this purpose as well.

In conclusion, we have shown in simulated outbreaks that, while vertical methods to analyze SWTs can preserve Type I Error and provide valid effect estimates, they are less powerful than parallel-arm CRT designs, which are themselves less powerful than IRT designs. Given the primary purpose of a randomized trial to demonstrate efficacy of the intervention, SWTs have serious statistical disadvantages compared to these other two designs.

## ACKNOWLEDGMENTS

This work was supported by the National Institute of Allergy and Infectious Diseases (Lee Kennedy-Shaffer, award number 1F31AI147745) and the National Institute of General Medical Sciences (Marc Lipsitch, award number U54GM088558).

The authors thank Dr. Matt Hitchings for making available his simulation code and Rebecca Kahn for support in using that code. The authors also wish to thank Dr. Victor De Gruttola and Dr. Michael D. Hughes for helpful comments on the methods and interpretation of results.

## Data Availability

Code to reproduce the simulations is available at https://github.com/leekshaffer/SW-CRT-outbreak.

## Notes

### Competing Interest Statement

The authors have declared no competing interest.

